# The infection evidence of SARS-COV-2 in ocular surface: a single-center cross-sectional study

**DOI:** 10.1101/2020.02.26.20027938

**Authors:** Xian Zhang, Xuhui Chen, Liwen Chen, Chaohua Deng, Xiaojing Zou, Weiyong Liu, Huimin Yu, Bo Chen, Xufang Sun

## Abstract

**Purpose:** The aim of this study was to identify whether SARS-COV-2 infected in ocular surface.

**Methods:** Cross-sectional study of patients presenting for who received a COVID-19 diagnosis, from December 30, 2019 to February 7, 2020, at Tongji hospital, Tongji medical college, Huazhong University of Science and Technology. Demographics, temperature was recorded, blood routine test (Rt), chest Computed Tomography (CT) were took intermittently, and SARS-COV-2 real-time reverse-transcriptase–polymerase-chain-reaction (RT-PCR) assay were arranged for the nasopharyngeal and conjunctival swab samples.

**Results:** A total of 102 patients (48 Male [50%] and 54 Female [50%]) with clinical symptoms, Rt, and chest Computed Tomography (CT) abnormalities were identified with a clinical diagnosis of COVID-19. Patients had a mean [SD] gestational age of 57.63 [14.90] years. Of a total of 102 patients identified, 72 patients (36 men [50%] and 36 women [50%]; mean [SD] age, 58.68 [14.81] years) confirmed by laboratory diagnosis with SARS-COV-2 RT-PCR assay. Only two patients (2.78%) with conjunctivitis was identified from 72 patients with a laboratory confirmed COVID-19. However, SARS-COV-2 RNA fragments was found in ocular discharges by SARS-COV-2 RT-PCR only in one patient with conjunctivitis.

**Conclusions:** Although we suspect the incidence of SARS-COV-2 infection through the ocular surface is extremely low, the nosocomial infection of SARS-CoV-2 through the eyes after occupational exposure is a potential route. The inefficient diagnostic method and the sampling time lag may contribute to the lower positive rate of conjunctival swab samples of SARS-COV-2. Therefore, to lower the SARS-COV-2 nosocomial infection, the protective goggles should be wore in all the health care workers.

## Introduction

A novel coronavirus, severe acute respiratory syndrome coronavirus 2 (SARS-COV-2) associated with severe human infected disease (COVID-19) outbroke starting from Wuhan city, in China.^1^ Most recently, Wang D et al reported 138 hospitalized SARS-COV-2 pneumonia cases with a hospital-associated transmission rate of 41%, among whom 70% were medical staffs.^2^ To date, nearly 80, 000 people have been infected by COVID-19, including over 3000 medical staffs. Therefore, in order to curb the spread of SARS-COV-2, and reduce the nosocomial infection, the exact routes of transmission need to be further studied and confirmed.

Although primarily affecting the respiratory tract, it also involves extra-pulmonary sites, including the digestive tract and other organs.^3^ SARS-CoV-2 infected patients develop respiratory illness, with the first symptoms of fever, cough and fatigue that quickly progress to pneumonia. COVID-19 were also contributed to extra-pulmonary manifestations in a number of patients at the onset of the illness, such as headache, diarrhoea, nausea and vomiting,^4^ or even presented with asymptomatic infection.^8^ Despite the intensive work that has been focused on an understanding of the transmission of SARS-CoV-2, the spread mode of the SARS-CoV-2 is unclear.

To our knowledge, limited investigations have been conducted to date into the clinical features of SARS-COV-2 in ocular surface. A few of COVID-19 patients with conjunctivitis, or even as the first symptom, make the early diagnosis and effective protection a great difficulty. A recent research has demonstrated that SARS-COV-2 can be detected in the conjunctival sac of patients with COVID-19 patients or suspected.^5^ However, none of these patients had ocular symptoms. We therefore conducted a study to describe the clinical spectrum of ocular symptoms and laboratory test in conjunctival swab samples, we found a rare case of nosocomial SARS-COV-2 infection with conjunctivitis in a nurse, which suggests that ocular transmission may be a potential route of nosocomial transmission of the SARS-COV-2.

## Methods

A Cross-sectional study was conducted of all patients who received a COVID-19 diagnosis between December 30 30, 2020, and February 7, 2020, at Tongji hospital, Tongji medical college, Huazhong University of Science and Technology. The aim of this study was to identify whether ocular surface is an infected target of SARS-CoV-2. This study was approved by the ethics committee of Tongji hospital and was performed in accordance with the tenets of the Declaration of Helsinki.^6^

Clinical specimens for 2019-nCoV diagnostic testing were obtained in accordance with CDC guidelines. To diagnosis the COVID-19, blood routine test (Rt), chest Computed Tomography (CT) and SARS-COV-2 real-time reverse-transcriptase–polymerase-chain-reaction (RT-PCR) assay were arranged. Serum was collected in a serum separator tube and then centrifuged in accordance with CDC guidelines. Laboratory-confirmed cases were identified with the criteria of at least one positive result from a respiratory specimen using RT-PCR assays. All patients invited to participate in the study provided consent for the nasopharyngeal and conjunctival swab samples with synthetic fiber swabs.

Then, these swab samples were detected via real-time RT-PCR assays as previously described.^7^ Briefly, RNA extraction from nasopharyngeal swabs and conjunctival swabs was performed RT-PCR on a Real Time PCR System. Thermal cycling was initiated with an initial incubation at 50°C for 15 min, 95°C for 3 min. Each PCR cycle consisted of heating at 95°C for 15 s for melting and at 60°C for 30 s for annealing and extension. The primers and probes for SARS-CoV-2 detection were selected according to the National Pathogen Resource Center (National Institute for Viral Disease Control and Prevention, China CDC), and the sequences were as follows^7^: forward primer 5′-ACTTCTTTTTCTTGCTTTCGTGGT-3′; reverse primer 5′-GCAGCAGTACGCACACAATC-3′; and the probe 5′CY5-CTAGTTACACTAGCCATCCTTACTGC-3′BHQ1.

The statistical analysis was performed using SPSS software version 22.0 (SPSS Inc, Chicago, IL). The descriptive data were presented as percentage, mean, and standard deviation. All the symptoms and examination results are present in the Powerpiont 2016.

## Results

A total of 102 patients (48 Male [50%] and 54 Female [50%]) with clinical symptoms, Rt, and CT abnormalities were identified with diagnosis of COVID-19. Patients had a mean [SD] gestational age of 57.63 [14.90] years. Of a total of 102 patients identified, 72 patients (36 men [50%] and 36 women [50%]; mean [SD] age, 58.68 [14.81] years) confirmed by laboratory diagnosis. The time for conjunctival sampling were had a mean [SD] day of 18.15 [7.57] days. Demographic data including patients’ age, gender and the time for conjunctival sampling were shown in **Fig. 1**.

**Fig.1.**
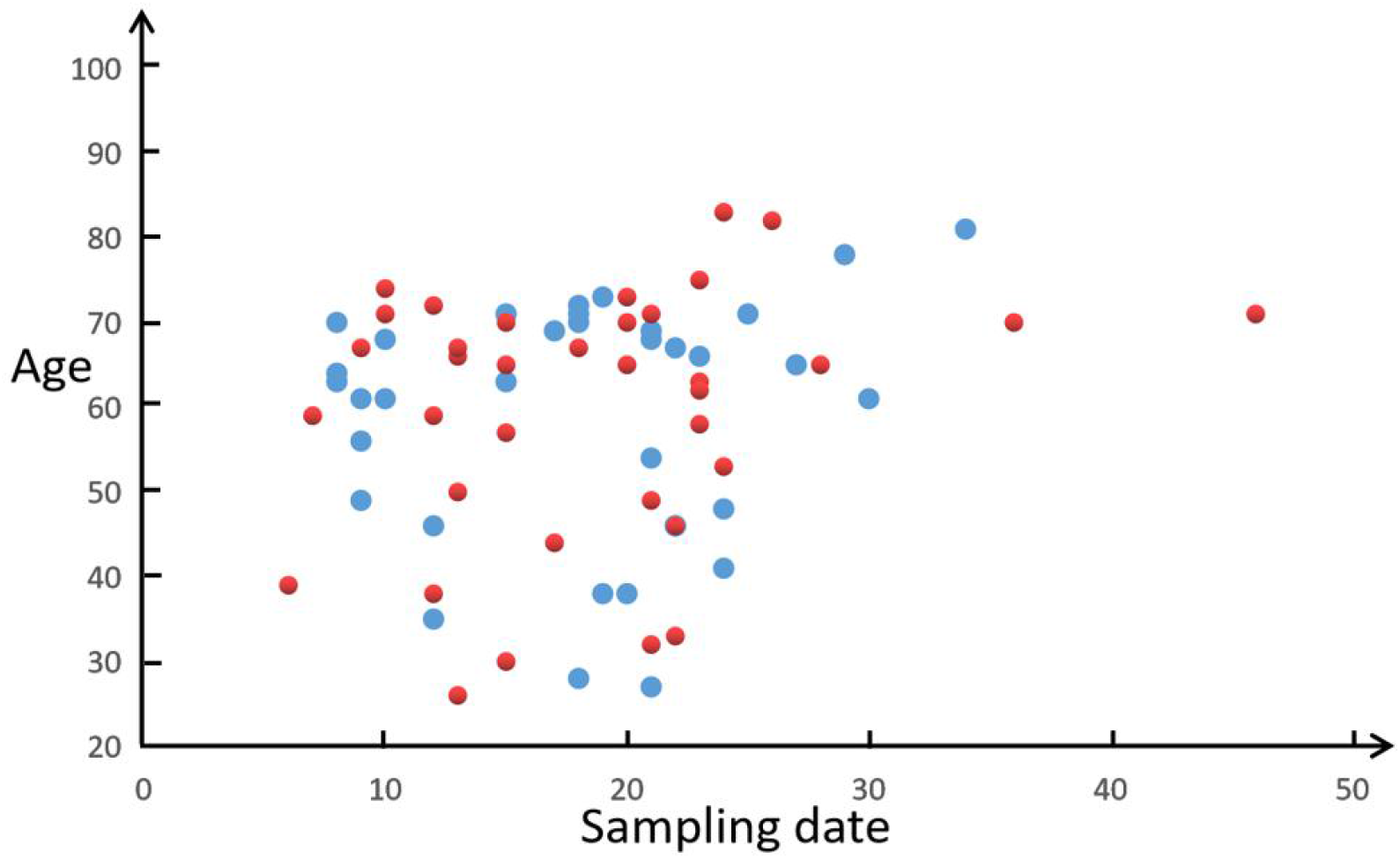
Demographic data: Scatter plot of the patients’ distribution characteristics according to the age, gender and sampling date from the date of onset. The sampling date of conjunctival swab varied from the 6th day to the 46th day, with an average of 18.15 days.(red spots: females; blue spots: males)

Only two patients (2.78%) with conjunctivitis were identified from 72 patients with a laboratory confirmed COVID-19. However, SARS-CoV-2 was found in ocular discharges by RT-PCR only in one patient, the symptoms and laboratory test were shown in **Fig. 2**. Briefly, a 29-year-old nurse working in the Emergency Department at Tongji hospital, Wuhan City, China was referred to the Department of Ophthalmology at Tongji Hospital on February 1st, 2020 due to excessive tearing and redness in both eyes (**Fig. 3A**). No other systemic symptoms except for a moderate fever of 38.2? was reported on January 31st, 2020. The ocular examination revealed conjunctival congestion and watery discharges in both eyes with normal best corrected visual acuity, normal corneal epithelium, quiescent anterior chamber and no tenderness or enlargement of the preauricular lymph node. We therefore excluded the possibility of conventional conjunctivitis, such as bacterial conjunctivitis, hemorrhagic conjunctivitis, allergic conjunctivitis, according to her clinical symptom and sign. She clarified medical N95 respirators continuously wore during operation, while occasionally worked with a dislocated eye mask touching her eyelids. Considering the occupational exposure by SARS-COV-2, SARS-COV-2 related assay were arranged. Surprising, the chest CT showed multiple peripheral ground-glass opacities in both lungs (**Fig. 3D-F**). The conjunctival and oropharyngeal swabs tested for SARS-COV-2 were both positive. The blood Rt showed a normal total white blood cells with elevated monocyte counts. On the basis of her epidemiologic characteristics, clinical manifestations, chest images, and laboratory findings, this patient was diagnosed with SARS-COV-2 infected acute viral conjunctivitis and pneumonia.

**Fig.2.**
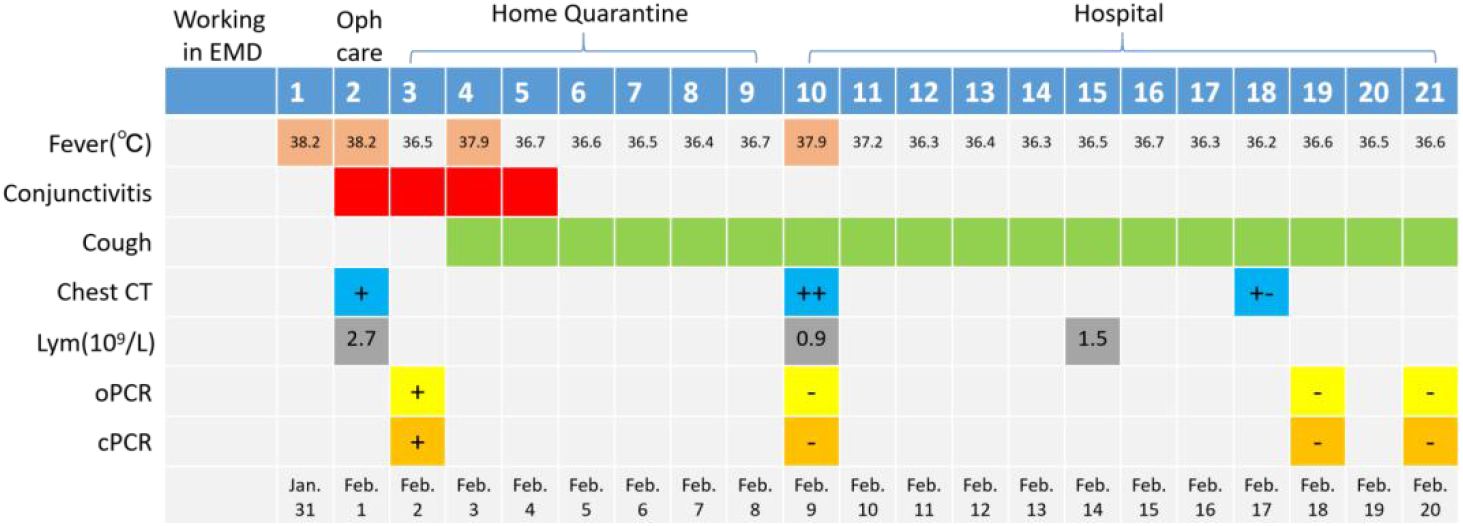
Maximum Body Temperatures, Symptoms, and Laboratory Tests of Positive Patient according to Day of Illness and Day of Hospitalization, January 31 to February 20, 2020. The early symtems were fever and conjunctivitis which last for 4 days. Both oropharyngeal and conjunctival swabs PCR were positive on the 3rd day followed by 3 negative tests. (oPCR: oropharyngeal swabs PCR; cPCR: conjunctival swabs PCR; Lym: Peripheral Blood Lymphocyte Count; EMD: Emergency Department; Oph: Ophthalmology Department.)

**Fig. 3.**
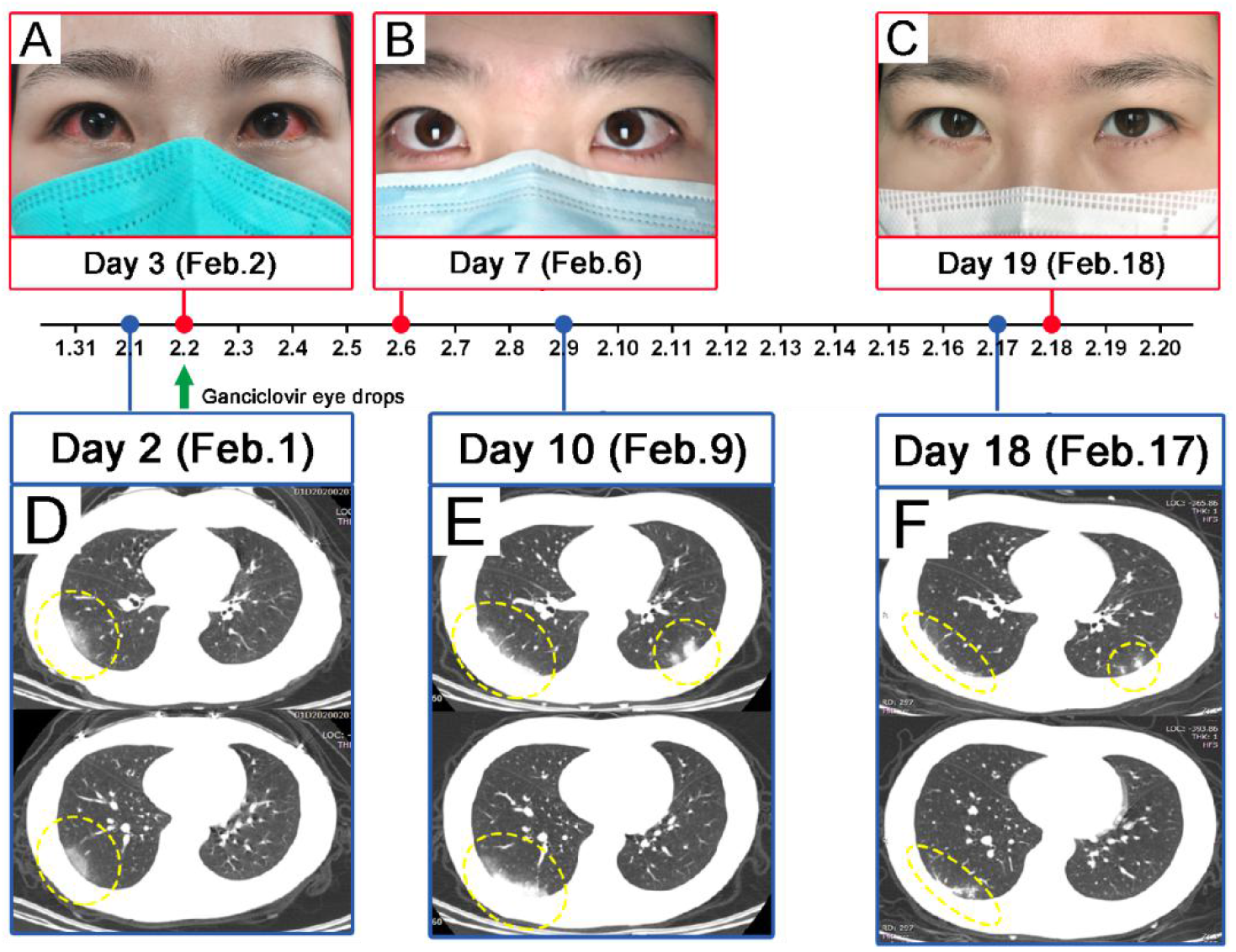
Clinical photographs of the positive patient regarding conjunctivitis and chest CT: (A) binocular conjunctival congestion which is a classic sign for viral conjunctivitis; (B,C) conjunctivitis completely subsided and did not recur 4 days after onset; (D-F) dynamic changes of peripheral ground-glass opacities in both lungs (yellow arrows) which illuminate the pneumonia caused by SARS-COV-2 infection.The pulmonary lesions were aggravated on the 10th day, and the lesions were basically absorbed on the 18th day.

Before admission, the patient reported persistent a 4-day history of conjunctivitis and a 3-day history of fever. Ganciclovir eye drops was used to control her conjunctivitis, the patient’s conjunctivitis vital signs remained stable during her home quarantine, apart from the development of intermittent fevers, accompanied by periods of cough from day 4 to 9 post of illness (**Fig. 2**). Although, SARS-COV-2 RT-PCR assay for the nasopharyngeal and conjunctival swab were negative 5 days after the conjunctivitis taken a turn for the better, a fever reappear, and the Chest Radiographs showed that her pneumonia aggravated at day 10 post illness. Then, she was arranged to the hospital treated according to the guidance of CDC, and then discharged 11 days post hospitalization, accompanied with a slight cough.

## Discussion

Like other highly contagious respiratory viruses, respiratory droplets are considered as the main route of SARS-COV-2 transmission.^8^ However, other possible transmission ways should be taken into account owing to the rapid transmission. A recent bioinformatics analysis study analyzed 4 datasets with single-cell transcriptomes of lung, oesophagus, gastric, ileum and colon, and SARS-CoV-2 was found in stool samples of patients with abdominal symptoms.^9^ Moreover, Lofy KH et al^10^ detected the SARS-COV2 in a patient’s stool, which warns us of the risk of fecal oral transmission. Similarly, SARS-CoV-2 was also directly detected in the oesophageal erosion and bleeding site in a case with severe peptic ulcer symptom reported by Zhong et al.^11^ Therefore, the real spread route is still a mystery, other transmitted ways should be further studied and confirmed.

In the current study, we found that two patients (2.78%) with conjunctivitis was identified from 72 patients with a laboratory confirmed COVID-19. However, SARS-CoV-2 was found in ocular discharges by RT-PCR only in one COVID-19 patient. Although the incidence of conjunctivitis is extremely low, these results demonstrated that SARS-CoV-2 have shown a capacity to use the eye as a portal of entry and cause ocular disease. To our knowledge, several anatomical and mucosal immune properties permitted the eye as both a potential site of virus infected site as well as a gateway for respiratory infection.^12-13^ In coincidence with our result, SARS-CoV was detected by RT-PCR in tear samples from three probable cases.^14^ Resemble as SARS-CoV, entry of SARS-CoV-2 via the host functional receptor is mediated by ACE2.^15^ Moreover, Sun and colleagues^16^ found that ACE2 expressed in human cornea and conjunctival tissues, which providing strong evidence for our diagnosis.

Furthermore, Dr. Guangfa Wang, a member of the national expert panel on pneumonia, reported that he was infected by SARS-COV-2 during the inspection in Wuhan, through unprotected eye exposure.^17^ Similarly, an anesthesiologist with insufficient eye protection confirmed with COVID-19 and also presented conjunctivitis as the initial symptom. The laboratory test revealed that the nasopharyngeal swab was positive while the conjunctival swab was negative.^5^ On the contrary, our case initially presented similar conjunctivitis with a positive conjunctival PCR result. Considering the same occupational exposure of Dr. Wang, the anesthesiologist and the nurse with positive conjunctival PCR result in our case, and the SARS-COV-2 infected mechanism, we believe that ocular transmission of SARS-CoV-2 must be seriously considered, especially in health care workers.

The negative results of conjunctival sac in other 71 patients, may be owing to the lower viral concentration, the sampling time lag, and the lower positive rate of the inefficient diagnostic method. Ziad and colleague^18^ found that tracheal aspirates yielded significantly higher SARS-CoV loads, compared with the nasopharyngeal swab and sputum specimens. This suggests that the viral concentration and genome fraction is diverse in different sites. In consideration of the ocular surface is an open microenvironment, and the viral may transport to the inferior meatus of the nose rapidly,^19^ the SARS-CoV-2 concentration in ocular surface is likely to be very low. We also found that suspected patients often had 2∼3 repeated tests of nasopharyngeal swabs before the positive result confirmed SARS-CoV-2 infection. The sampling time lag and the inefficient diagnostic method may contribute to this lower positive rate of SARS-CoV-2. de Wit and colleagues^20^ demonstrated that, in the rhesus macaque model, MERS-CoV RNA could detect in the conjunctiva, and the viral loads could no longer be detected in the conjunctiva 6 days post infection. However, the mean time for conjunctival sampling are 18.15 days in our present study. Another thing needs to be considered is that the lower positive rate of RT-PCR makes early diagnosis of SARS-CoV-2 a challenge. Therefore, improvements in the sensitivity of molecular diagnostic methods need to be taking in the future.

There are several limitations need to be considered. First, the conjunctival scraping test should be done as early as possible if we find ocular symptoms in a suspected one. The mean time for conjunctival sampling in our study are 18.15 days, in which exceeded the optimal detection time. Second, how to increase the positive rate. The most commonly used chest CT examination has certain limitations for the special suspected groups, such as pregnant women. In addition, we found that 2∼3 repeated tests did in suspected patients of nasopharyngeal swabs before the SARS-CoV-2 was obtained, which may be related to the false-negative result of RT-PCR. Therefore, the low viral load in the ocular surface and the lower positive rate of RT-PCR makes early diagnosis of SARS-CoV-2 a challenge. Recently, Doan^21^ reported that the influenza virus and rubella virus found in patients’ conjunctival sac or tears by the next generation sequencing rapidly, which provides us with a feasible direction for future study.

## Conclusions

In conclusion, we suspect the incidence of SARS-COV-2 infection through the ocular surface is extremely low in the general population, however, considering the common feature of the positive cases which are occupational exposure accompanied with conjunctivitis in the early stage, the higher viral aerosol load in the hospital and more opportunities to contact with COVID-19 patients, we highlight that ocular transmission is a potential important way of occupational exposure for medical staff. Therefore, To lower the risk of SARS-COV-2 nosocomial infection, the protective goggles should be wore in all the health care workers, especially who work in the Fever Outpatient and Infection Wards. Patients with conjunctivitis in the epidemic area should also be treated seriously to rule out the COVID-19. Moreover, to date, the lower positive rate of conjunctival sac may be ascribed to the lower viral concentration, the sampling time lag, and the inefficient detection methods. Our study highlights the need for further research into the efficient detection methods of SARS-COV-2.

## Data Availability

Data sharing not applicable to this article as no datasets were generated or analysed during the current study.

